# Global COVID-19 fatality analysis reveals Hubei-like countries potentially with severe outbreaks

**DOI:** 10.1101/2020.03.26.20038075

**Authors:** Boyan Lv, Zhongyan Li, Yajuan Chen, Cheng Long, Xinmiao Fu

**Affiliations:** Provincial University Key Laboratory of Cellular Stress Response and Metabolic Regulation, College of Life Sciences, Fujian Normal University, Fuzhou City, Fujian Province 350117, China; Department of Orthopaedic, Sichuan University West China Hospital, Chengdu City, Sichuan Province, China

**Keywords:** COVID-19, SARS-CoV-2, coronavirus, epidemic, crude fatality ratio, mortality

## Abstract

1. CFR in Iran in the early stage of the outbreak is the highest among all the countries 2. CFRs in the USA and Italy are similar to that in Hubei Province in the early stage of the outbreak. 3. CFRs in South Korea are similar to that outside Hubei (in China), indicating less severe outbreaks therein. 4. Our findings highlight the potential severity of outbreaks globally, particular in the USA.

---

The outbreak of 2019 novel coronavirus diseases (COVID-19) is ongoing in China ^1^, but appears to reach late stage and also just starts to devastate other countries ^2^. As of 13 March 2020, there have been 80991 confirmed COVID-19 cases and 3180 deaths in China, much higher than those outside China with 51767 confirmed cases and 1775 deaths ^3^. However, the daily increase in COVID-19 cases outside China has greatly surpassed that inside China ^3^ (over 7000 verse 11 on 13 March), and therefore people raise deep concerns about the outbreaks outside China. Here we attempted to uncover their characteristics by comparative analysis on crude fatality ratios (CFRs).

We collected data on the officially released cumulative numbers of confirmed cases and deaths (from 23 January to 13 March 2020) with respect to mainland China, epicenter of the outbreak (i.e., Hubei Province and Wuhan City), outside Hubei (in China) and outside Wuhan (in Hubei), as well as to typical countries reported with a substantial number of deaths including South Korea, Japan, Iran, Italy, USA, France and Spain (**Fig. 1**). CFRs in Hubei and Wuhan are significantly higher than those outside Hubei and outside Wuhan, and they are relatively higher in the early stage of outbreaks than in the late stage (**Fig. 1A**), in line with earlier comprehensive reports by China CDC and WHO ^4, 5^. The outbreaks outside China are overall lagging approximately one month behind China (**Fig. 1B** vs **Fig. 1A**). CFR in Iran in the early stage (from 21 February to late March) is extremely high while CFR in Korea is low and stable over time. Notably, CFR in Iran has significantly decreased since 2 March while CFR in Italy increased a lot in the past 10 days.

**Figure 1.**
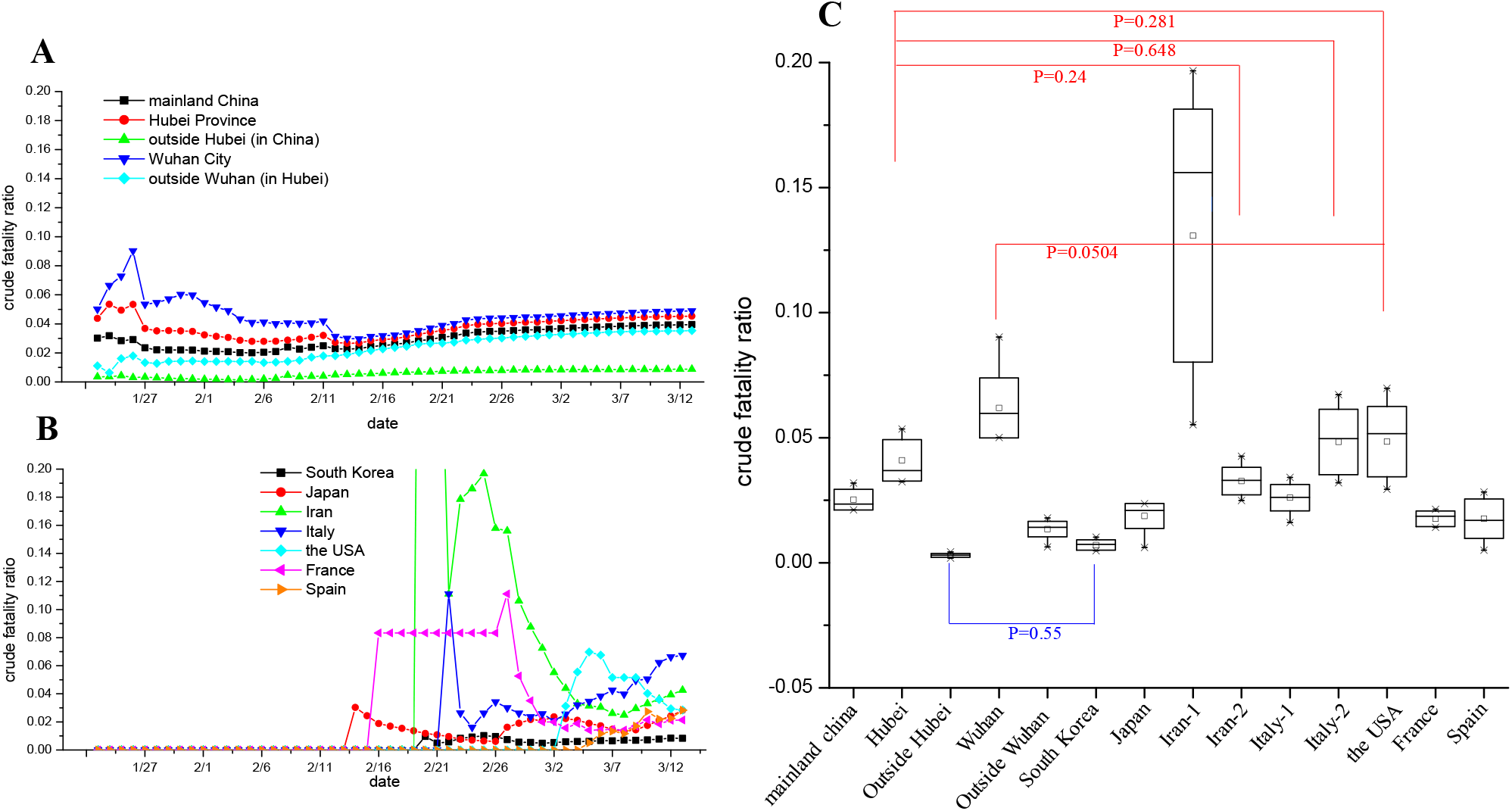
CFR comparisons between China and outside China. (**A, B**) CFR for COVID-19 in the indicated regions of China (panel A) or in the indicated countries (panel B) over time (from 23 January to 13 March 2020). (**C**) Difference analysis of CFRs in the early stages of COVID-19 outbreaks between China and outside China. CFRs in a period of 10-day, i.e., from 23 March to 1 February for China and other specific periods for countries outside China (for detail, refer to S1.xls file), were plotted as mean±SD at 95% confidence intervals (in the black box), with median being shown as short lines. Statistics were performed using SPSS with ANOVA algorithm, and significance levels (*P* value) for all the pairs are shown in **Table S1**. P values larger than 0.05 between Wuhan/Hubei and other countries are colored in red, indicating no significant difference (i.e., somehow being similar to each other) and the relative severity of the epidemic therein; P value between outside Hubei and South Korea is 0.55 (colored in blue), indicating relatively mild or controllable epidemic in South Korea.

Next, we performed statistical analysis on CFRs in a period of 10 days in the early stage of outbreaks between outside China and China. In particular, two periods were set for Iran and Italy in order to fully cover their changing trends (for detail, refer to S1.xls file). Results displayed in **Fig. 1C** revealed i) CFRs in Iran, Italy and USA in the past ten are not significantly different from Hubei (*P* being 0.24, 0.648 and 0.281, respectively); ii) CFR in USA is not significantly different from Wuhan to marginal degree (*P* being 0.0504); iii) CFR in Iran from 22 February to 2 March is significantly different from any regions of China (p<0.001; **Table S1**). In view of the detailed *P* values among all pairs (**Table S1**), we suppose the ranking for the severity of COVID-19 outbreaks in different countries/regions in terms of CFRs as follows: Iran>Wuhan>Hubei≈USA≈Italy>outside Wuhan ≈Spain≈Japan≈France>South Korea≈outside Hubei.

As CFR is defined as the number of deaths (numerator) among the number of confirmed cases (denominator), both increase of numerator and decrease of denominator lead to higher CFR. In Hubei/Wuhan there were neither sufficient COVID-19 test kits for infection identification nor enough beds in hospitals for effective treatments in the early stage of the outbreak ^6^. These shortages led to numerous transmissions in households, reduced the apparent number of cumulative confirmed cases and caused mild patients without treatments to become severe/critical ones and even die, as implicated by earlier reports ^4, 7^. As such, CFRs in Hubei/Wuhan was relatively high in the early stage ^5, 7^. Similar CFRs between Hubei and USA/Italy, suggest that these countries may face similar situations at present as Hubei had experienced before. In support of this, recent news reports show that Italy is extremely short of medica resources (beds and acute care equipment) while USA has some problems in COVID-19 test ^8^. In Iran, these problems might be even more severe such that its CFR is extremely high. To fight against the COVDI-19 outbreaks in these Hubei/Wuhan-like countries, governments may need to implement control measures and timely supply medical resources as Hubei/Wuhan had done in the past month ^2, 4^.

## Data Availability

all included in the manusccript

## Acknowledgments

This work is support by the National Natural Science Foundation of China (No. 31972918 and 31770830 to XF). Authors declare no conflict of interests.

## Supporting information file

### Methods

#### Sources of data

We collected the cumulative number of confirmed cases and deaths (from 23 Jan 2020 to 7 March 2020) of COVID-19 from the official websites of the National Health Commission of China (http://www.nhc.gov.cn/xcs/yqtb/list_gzbd.shtml) and Hubei Provincial Health Commissions (http://wjw.hubei.gov.cn/fbjd/dtyw/) and WHO’s website (https://www.who.int/emergencies/diseases/novel-coronavirus-2019/situation-reports/). Crude fatality ratio was calculated as the cumulative number of COVID-19 deaths among the cumulative number of confirmed cases with respect to each region or country.

It should be pointed out that for USA, only five-day data points are available because its first COVID-19 death was reported on 3 March 2020. We noticed that: i) In the WHO Situation Report-48 (i.e., on 8 March), the data of USA are the same with those on 7 March [1], apparently no update; ii) The data of USA from the real-time COVID-19 website (https://news.sina.cn/project/fy2020/yq_province.shtml?country=SCUS0001&wm=6109&wm=6109) of Sina, a top internet corporation in China, are substantially different from the data released by WHO [2] (for detail, refer to **Table S2**), but consistent with the data from Baidu, another top internet corporation in China (https://voice.baidu.com/act/newpneumonia/newpneumonia/?from=osari_pc_3#tab4). In view of these, the data for USA were also downloaded from Sina and analyzed.

### Results

**Table S1.** Significance levels for CFR comparisons between different geographic regions.

|  | China | Hubei | outside<br>Hubei | Wuhan | outside<br>Wuhan | South<br>Korea | Japan | Iran-1 <sup>b</sup> | Iran-2 <sup>b</sup> | Italy-1<br><sup>b</sup> | Italy-2<br><sup>b</sup> | USA | France | Spain |
| --- | --- | --- | --- | --- | --- | --- | --- | --- | --- | --- | --- | --- | --- | --- |
| China |  | 0.024 | 0.002 | 0 | 0.091 | 0.01 | 0.349 | 0 | 0.274 | 0.908 | 0.007 | 0.001 | 0.231 | 0.155 |
| Hubei | 0.024 |  | 0 | 0.003 | 0 | 0 | 0.002 | 0* | 0.24 | 0.032 | 0.648 | 0.281 | 0.001 | 0 |
| outside<br>Hubei | 0.002 | 0 |  | 0 | 0.131 | 0.55 | 0.024 | 0 | 0 | 0.001 | 0 | 0 | 0.046 | 0.091 |
| Wuhan | 0 | 0.003 | 0 |  | 0 | 0 | 0 | 0* | 0 | 0 | 0.011 | 0.054 | 0 | 0 |
| outside<br>Wuhan | 0.091 | 0 | 0.131 | 0 |  | 0.36 | 0.448 | 0 | 0.006 | 0.071 | 0 | 0 | 0.62 | 0.823 |
| South<br>Korea | 0.01 | 0 | 0.55 | 0 | 0.36 |  |  | 0 | 0 | 0.007 | 0 | 0 | 0.159 | 0.265 |
| Japan | 0.349 | 0.002 | 0.024 | 0 | 0.448 | 0.095 | 0.095 |  | 0.044 | 0.293 | 0 | 0 | 0.792 | 0.607 |
| Iran-1 | 0 | 0 | 0 | 0 | 0 | 0 | 0 | 0 |  | 0 | 0 | 0 | 0 | 0 |
| Iran-2 | 0.274 | 0.24 | 0 | 0 | 0.006 | 0 | 0.044 | 0 | 0 |  | 0.104 | 0.025 | 0.023 | 0.014 |
| Italy-1 | 0.908 | 0.032 | 0.001 | 0 | 0.071 | 0.007 | 0.293 | 0 | 0.328 | 0.328 |  | 0.001 | 0.189 | 0.125 |
| Italy-2 | 0.007 | 0.648 | 0 | 0.011 | 0 | 0 | 0 | 0 | 0.104 | 0.01 | 0.01 |  | 0 | 0 |
| USA | 0.001 | 0.281 | 0 | 0.054 | 0 | 0 | 0 | 0 | 0.025 | 0.001 | 0.533 | 0.533 | 0 | 0 |
| France | 0.231 | 0.001 | 0.046 | 0 | 0.62 | 0.159 | 0.792 | 0 | 0.023 | 0.189 | 0 | 0 |  | 0.796 |
| Spain | 0.155 | 0 | 0.091 | 0 | 0.823 | 0.265 | 0.607 | 0 | 0.014 | 0.125 | 0 | 0 | 0.796 |  |

<sup>a</sup> P values between countries and Hubei/Wuhan, if >0.05, indicate relative severe outbreaks therein and are colored in red; P values between countries and outside Hubei, if >0.05, indicate less severe outbreaks therein and are colored in blue; P values between Hubei/Wuhan and Iran-1 is less than 0.001 (colored in purple, indicating the extremely severe outbreak).
<sup>a</sup> Two sets of data for Iran and Italy in different periods were analyzed (for detail, refer to Table S2).

---

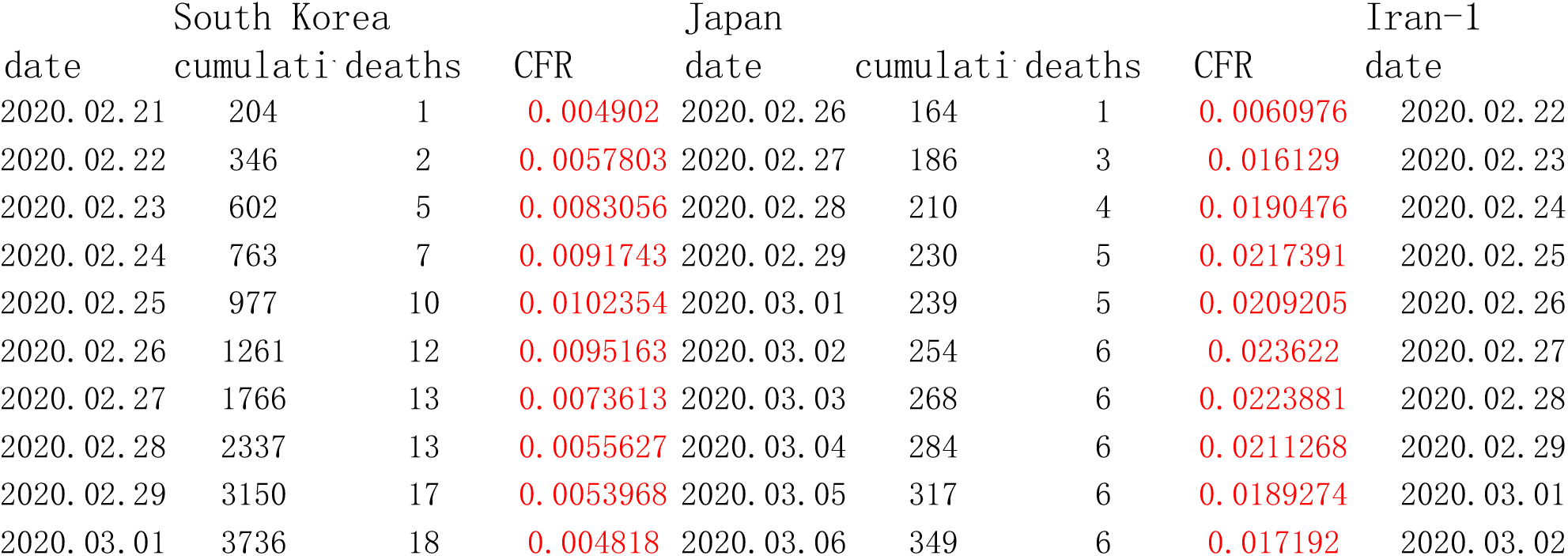

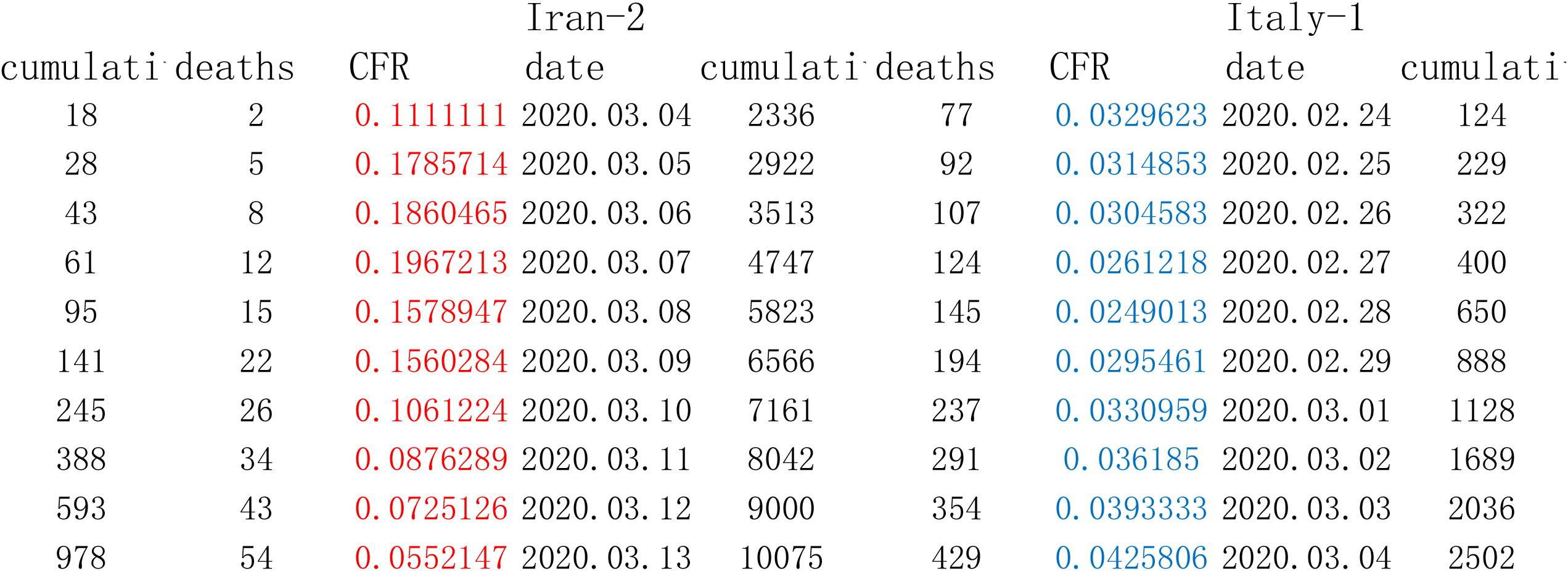

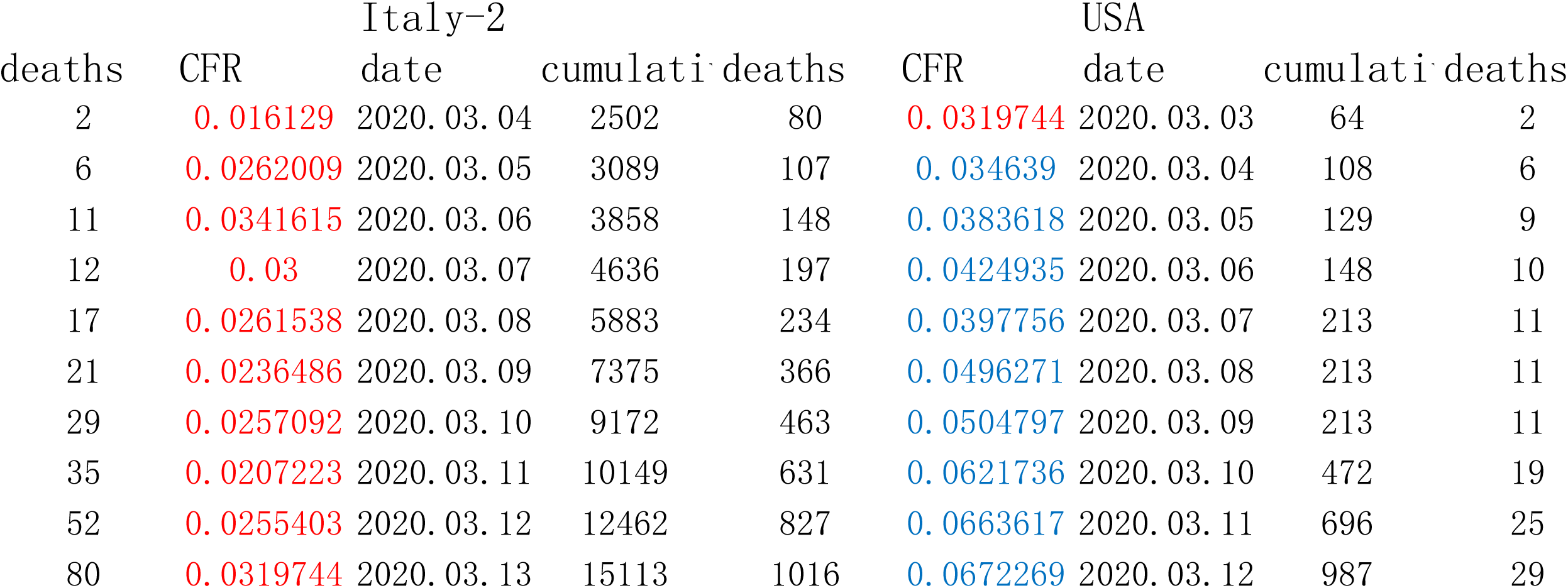

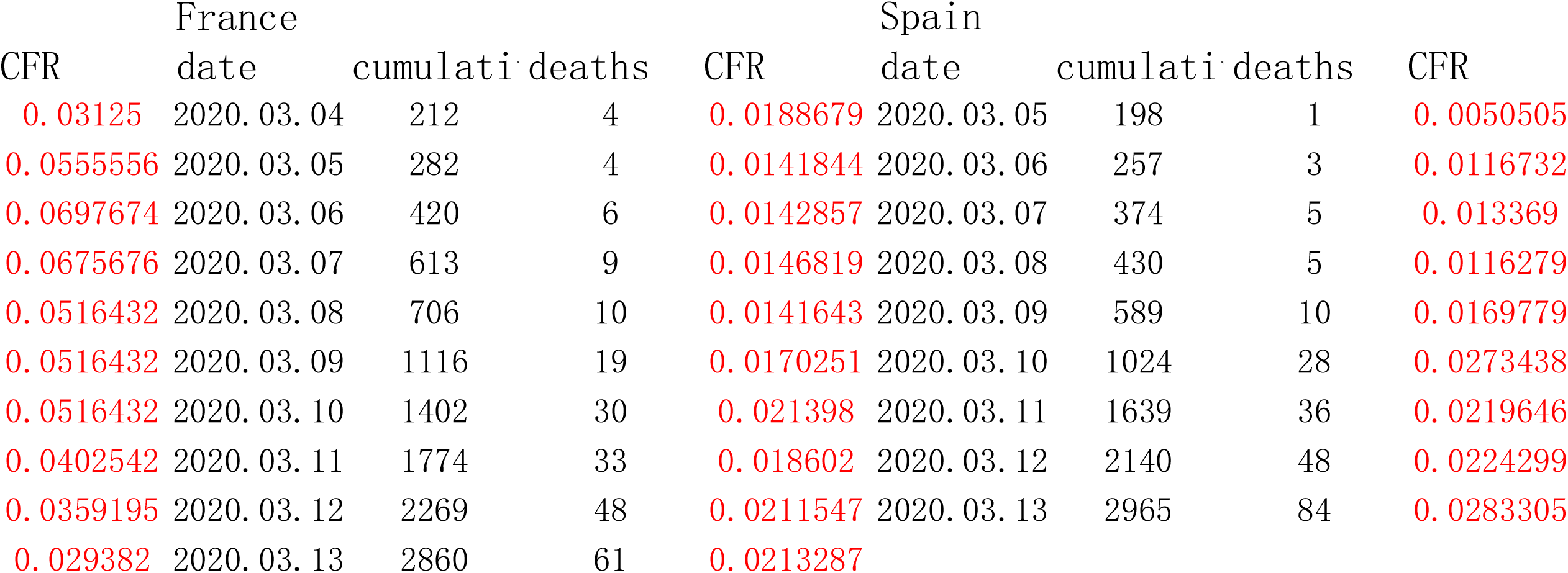

## Notes

### Competing Interest Statement

The authors have declared no competing interest.

